# Job insecurity, financial threat and mental health in the COVID-19 context: The buffer role of perceived social support

**DOI:** 10.1101/2020.07.31.20165910

**Authors:** Carlos-María Alcover, Sergio Salgado, Gabriela Nazar, Raúl Ramírez-Vielma, Carolina González-Suhr

**Affiliations:** Departamento de Psicología, Universidad Rey Juan Carlos, Madrid, Comunidad de Madrid, España; Departamento de Administración y Economía, Universidad de La Frontera, Temuco, Región de la Araucania, Chile; Departamento de Psicología, Universidad de Concepción, Concepción, Región del Biobío, Chile

## Abstract

The social distancing, confinement and quarantine adopted since March 2020 to confront the COVID-19 pandemic have affected multiple vital areas, and specially work, business and productive activities. Prior research has highlighted the relation between perceptions of risk in employment and its concomitant financial risk with a myriad of consequences for people’s well-being and health. In order to analyze the potential negative consequences of temporary layoffs, downsizing or closure of companies and businesses, and the consequent insecurity about the continuity of employment, the aim of this study is twofold. Firstly, to analyze the relations between the perceptions of job insecurity and financial threat and overall mental health during the first month of the COVID-19 pandemic in a sample of the Chilean adult population. And secondly, to identify the potential buffer effect of perceived social support on this relation. To analyze this, we carried out a cross-sectional study on a non-probabilistic sample aimed at a general Chilean adult population. The results show that both perceptions of job insecurity and financial threat are associated with a decline in perceived mental health. Additionally, results indicate a moderate buffer effect of perceived social support relative to the size of the social network. Thus, in relation to job insecurity and financial threat, the network size mitigates the association of both with the decline in perceived mental health. The theoretical and applied scope of these findings are analyzed, and their challenges and limitations are discussed.

## Introduction

The disruptive effects of social distancing, confinement and quarantine adopted to confront the Covid-19 pandemic have affected multiple vital areas. Since March 2020, with its rapid expansion in Europe and later in America, work activities have suffered drastic changes. On the one hand, governments and businesses imposed or recommended working from home or remotely, which affected all types of activities, except for the essential sectors and services where being present was unavoidable (hospitals, basic product sourcing, pharmacies, food sales, etc.). On the other hand, a good number of productive sectors suddenly suspended their activities, affecting the majority of commerce, small services and hospitality businesses as well as industry in other activities considered non-essential, which resulted in the loss or temporary layoffs for a high number of workers. The economic implications of the global pandemic, called ‘Coronanomics’ [1], are still difficult to estimate in all their macro and micro magnitude on a global scale and per country [2]; however, the effects on people and their families who lost their job, suffered a temporary layoff or have kept it but perceive its possible loss or a deterioration in their working conditions, are already evident and can be identified.

Research conducted in the two last decades, and particularly since the beginning of the Great Recession of 2008, has uncovered the relation between perceptions of risk in employment and its concomitant financial risk with a myriad of consequences for people’s well-being and health. Thus, job insecurity (e.g. [3-5]) and financial insecurity (e.g. [6-8]) have negative effects on many aspects of people’s physical, mental and psychosocial health, including even direct effects on mortality when health is fragile [9] or it leads to suicidal behaviors [10], as well as affects family [11] and partner relationships.

On the other hand, previous research has shown the buffer effect of social support on the negative effects of job insecurity [12, 13]. Thus, the support received at work mitigates the negative effect of job insecurity on certain work-related outcomes, such as job dissatisfaction and noncompliant job behaviors, whereas the support received outside of work does so in relation to life dissatisfaction [14]. There is also empirical evidence of the buffer role of tangible social support in the relation between financial stress and psychological well-being and psychosomatic disorders [15], an effect that has been verified for both instrumental and emotional support [16].

In Chile, where this study was conducted, social distancing and confinement were adopted in mid-March 2020, which meant shutting down and closing multiple productive activities with the resulting temporary layoff or suspension of the work contract for a high number of workers, as well as the increase in uncertainty about future job continuity for many employees in vulnerable work situations. In order to analyze the potential negative consequences of these experiences, the aim of this study is twofold. Firstly, to analyze the relations between the perceptions of job insecurity and financial threat and overall mental health during the first month of the pandemic in a sample of the Chilean adult population. And secondly, to identify the potential buffer effect of perceived social support on this relation. Thus, our goal is carrying out an initial analysis of how work and economic factors can affect people’s mental health in a context of a prolonged health emergency and the concomitant economic crisis.

## Theoretical Background and Hypothesis

### Consequences of Job Insecurity on Mental Health and Health Correlates

Job insecurity refers to “the perceived threat of job loss and the worries related to that threat” [17, p. 1]. Basically, it is a subjective anticipatory perception, the core of which is concern and fear regarding the future continuity of one’s current job in the short or medium term, i.e., of involuntarily losing one’s job with all the related negative consequences for well-being and mental health, job attitudes and behaviors, and quality of life [18-20]. Job insecurity can be experienced in two ways, both in the sense of full job loss or quantitative insecurity [18, 21], and by changes in what, how, where and when work is done, or job status insecurity, “relating to anxiety about changes to valued features of the job” [22, p. 36], also called qualitative insecurity [21].

In short, the characteristics of job insecurity are [23]: subjective experience, future-focused phenomenon, perception, and response to anticipated visualization of (job or job valued features) loss. Further, experiences of job insecurity depend on three threat features: perceived situational control, threat duration and volition. Consequently [23], lower control, longer duration and lower volition will cause increased distress in workers, with detrimental effects on personal physical and psychological health and work-related well-being [5, 23-24]. Finally, other authors [26] have proposed differentiating the cognitive components of job insecurity related to the perception of loss or negative job change, from the affective components related to the emotional reactions to job loss or potential job change. The meta-analytic results [26] show that affective job insecurity is more significantly related to most of the correlates and outcomes than cognitive job insecurity. However, most research has used a global perspective (i.e., one-dimensional) of quantitative job insecurity (e.g. [18, 20, 27]), and this is the approach and measure adopted in this study.

Extensive prior research has identified the most relevant effects of job insecurity. First, the meta-analytic results had shown an overall negative effect on general mental health [19, 28], as well as for specific disorders such as anxiety, depression, decrease on psychological well-being, emotional exhaustion or life dissatisfaction [29]. Job insecurity is also associated with increased anxiety and irrational thoughts as well as psychological distancing at work, similar to what is experienced when face death or dismemberment [30]. Furthermore, negative effects of job insecurity have also been found on physical health, including headaches or eyestrain and skin problems [31], and incident coronary heart disease [32].

Second, it has also been found that the negative effects of job insecurity on health indicators and life satisfaction are greater in workers with low levels of employability or lack of opportunities to find a new job in case of losing the current one [31]. These effects are compounded when people are over 40 years of age [33].

Finally, an indirect effect of job insecurity on health can occur when people reduce their investments in health so they can save in order to cope with a possible job loss. Although this potential effect could only be verified in the long term [31], it is important to consider it as a factor that increases the accumulated vulnerability of workers with fewer resources, since there is evidence [34] that job insecurity also affects negatively daily consumption and some major life decisions.

> Based on these rationales and prior data, we formulate the following hypothesis:
>
> H1: Perceived job insecurity as a result of the COVID-19 pandemic will have a negative effect on overall mental health.

### Consequences of Financial Threat on Mental Health and Health Correlates

Financial threat is an emotional state which refers to “self-reported fearful-anxious uncertainty regarding one’s current and future financial situation” [35, p. 129; see also [7]. Financial threat perception, or financial distress, is usually associated with people or households that experience job insecurity when their jobs are less secure and have low protection against unemployment [36], and the data confirm that the financial threat is higher for people who experience a large number of economic difficulties in their daily life [37]. Economic difficulties can also become a chronic stress situation in families which can generate distressing thoughts about paying household expenses, and leads to feelings of fear, anxiety and uncertainty regarding one’s ability to maintain the current standard of living [7, 35]. Financial stress frequently forces high levels of debt to be incurred through credit cards, loans and mortgages [35]. There is evidence [38] that the negative effect of job insecurity on perceived health worsens if the person is over-indebted, Moreover, having non-mortgage debts (i.e., bank cards, loans or medical bills) cause higher levels of anxiety and damage to health than mortgages [38, 39].

Prior research has found that economic insecurity is a socioeconomic determinant of mental health [40]. Financial threat, preceded or not by debts already incurred, is associated with greater psychological distress, characterized by higher levels of depression, anxiety and fatigue [35], lower subjective well-being [41], greater emotional exhaustion and lower psychological well-being [37], and psychological distress and mental health issues [7]. Additionally, it has been proven that the perception of future financial risk (without future unemployment necessarily occurring) affects more negatively mental health than real volatility, which also affects all income levels and is more detrimental for men [40]. Worry and rumination about financial risk can also exacerbate their negative consequences on mental health, psychological well-being and cognitive functioning [42]. It has also been found that the perception of financial risk mediates the relation between economic difficulties and suicidal thoughts and cognitive confusion/bewilderment [43], affecting women and men equally.

The combination of job insecurity and financial insecurity or risk has a significant effect on lower levels of vitality and mental health [8], and it has been demonstrated [44] that in situations of economic turbulence (such as great crises and recessions, or like the one now caused by the COVID-19 pandemic), workers perceive a loss of the sense of control [45], so that the negative effects of job insecurity are even greater on mental health and perceived happiness.

> Based on these rationales and prior data, we formulate the following hypothesis:
>
> H2: Perceived financial threat as a result of the COVID-19 pandemic will have a negative effect on overall mental health.

### Perceived Social Support as Buffering in the Relationship between Job Insecurity, Financial Threat and Mental Health

Prior research has collected data and solid arguments to show a direct association between social relations, social support and health [46, 47]. In particular, the hypothesis of the buffer (protective) effect of the perception of social support on the consequences of stress has a long trajectory in psychosocial research. In their classic study, [48] found support for the buffer model when social support refers to the perceived availability of interpersonal resources that respond directly to the needs of people who are experiencing stressful events. Given that both job insecurity and financial threats constitute powerful work-related stressors [7, 17-18, 35], identifying the potential moderating effect of the perceived social support from a person’s social network is of great relevance to offsetting the negative consequences of both for health.

Theoretical models [30] and previous studies have suggested and demonstrated that social support buffers the negative effects of job insecurity on work outcomes, such as job satisfaction and vigor [49], and on well-being and mental health [12-14]. It has also been found that women use this social resource more often and more efficiently than men [50]. These results were confirmed in a recent study [51] that showed when women actively resort to their social support network, this acts as a buffer for the negative consequences of job insecurity. These conclusions agree with those proposed by [48] that the effectiveness of this interpersonal-social resource depends on it serving specifically to manage the stressful situation. However, another recent study [52] clarified previous results that supported the buffer effect of the perception of social support, as they showed that the negative relation between job insecurity and psychological well-being is reduced when the perception of job insecurity is low, but not when it is high.

As already we mentioned, there is also empirical evidence of the buffer role of tangible social support in the relation between financial stress and psychological well-being and psychosomatic disorders [15], and in the relation between financial stress and alcohol-related behaviors (drinking to deal with problems, excessive consumption of alcohol and problems with alcohol) [53]. The buffer effect has been verified for both instrumental and emotional social support [16]. These last results lead to the assumption that instrumental support can serve as a strategy centered on confronting the problem, whereas emotional support can be used as strategy centered on negative emotional responses, relations that may be of importance in the design of interventions to reduce to the negative effects of insecurity and financial threats.

> Based on these rationales and prior data, we formulate the following hypothesis:
>
> H3: The perception of social support linked to the size of the social network will mitigate the relations between perceived job insecurity and financial threat as a result of the COVID-19 pandemic, and overall mental health.

## Materials and Methods

The proposed hypotheses involve contrasting the existence of a relation between the variables in the context of the COVID-19 pandemic. No questions are considered about the point at which the relation between the variables begins or how long they last. Consequently, to prove the hypotheses, a cross-sectional correlational study was conducted on a non-probabilistic sample aimed at a general Chilean adult population. The project underwent assessment by the Ethics and Bioethics Committee of the Universidad de Concepción (CEBB 650-2020, March 2020), and the participants were asked to sign an informed consent.

### Participants

A sample of 591 participants took part in the study (592 responses to the online questionnaire were obtained, of which a duplicate case identified through e-mail was eliminated) with an average age of 37.63 years (*SD* = 12.85). 76 % were female (*n* = 449), 23.7% were male (*n* = 140), and 0.3% identified as another gender (*n* = 2). All the participants lived in Chile, with the Regions of Bio-Bio (*n* = 213; 36%), Metropolitan (*n* = 137; 23.2%), La Araucanía (*n* = 121; 20.5%), Valparaíso (*n* = 36; 6.1%), and Los Lagos (*n* = 17; 2.9%) being the most represented. Most of the participants during the study had a full-time day job (*n* = 269; 45.5%), but they also had part-time jobs (*n* = 43; 7.3%), were independent workers (*n* = 82; 13.9%), students (*n* = 79; 13.4%), unemployed (*n* = 55; 9.3%), retired (*n* = 30; 5.1%), homemakers (*n* = 24; 4.1%) and others (*n* = 9; 1.5%). 68.2% (*n* = 403) had a partner during the study. Participants had an average of 1.28 children (*SD* = 9.74). 93.1% lived in an urban area (*n* = 550) and 6.9% in the country (*n* = 41). 37.6% (*n* = 222) reported having graduate studies, 43.3% (*n* = 256) undergraduate studies, 8.3% (*n* = 49) technical-professional studies, 10.7% (*n* = 63) had high school and 0.2% (*n* = 1) elementary school.

### Procedure

The Government of Chile began to decree social distancing and confinement measures on March 16, 2020, and they progressively increased restrictions on productive, commercial and consumption activities as different regions detected contagions and deaths. The data were collected via an online questionnaire (Questionpro) between March 24 and April 23. Our recruitment strategy was a combination of professional networks, social networks, professional associations, undergraduate and graduate students and personal contacts, who were invited to participate and disseminate this invitation among their network of contacts (snowball strategy). When the proposed conditions were agreed upon, the participants marked a square indicating they were over 18 years, had been informed and understood the nature of this study and their decision to participate voluntarily. The battery of questionnaires included other instruments not considered in the present work as they deal with a complementary line of enquiry. The participants took an average of 27 minutes to complete all the instruments. When necessary, they could save their progress and take it up again when convenient.

### Instruments

#### Job Insecurity

To measure the perception of job insecurity, the Job Insecurity Scale developed by [27] was used, adapted and validated in Spanish [20] that includes four items (e.g., “I fear that I might lose my job”) scored on a 6-point Likert scale (1 = Strongly disagree, to 6 = Strongly agree). The instrument showed good internal consistency (Cronbach’s *alpha* = 80.1).

#### Financial Threat

The perception of financial threat was measured with the Financial Threat Scale adapted to Spanish [37]. This instrument contains six items (e.g., “How much do you feel threatened by your financial situation?”) that are evaluated on a 5-point Likert scale (1 = *Not at all*; 5 = A great deal). The scale showed good internal consistency (Cronbach’s *alpha* = 91.8).

#### Overall mental health

To measure the participants’ perception of overall mental health, the 12-item version of the General Health Questionnaire was used [54] in its version previously applied in Chile [55, 56]. This instrument is widely used as mental health screening [57]. It contains 12 questions (e.g., “Are you able to concentrate on what you do?”, “Have you felt constantly under strain?”, “Have you felt unhappy or depressed”) which is answered on a 4-point Likert scale (1 = *Never*; 2 = *Sometimes*; 3 = *Often*; 4 = *Always*), where high scores express greater decline in perceived overall mental health. Although at times in the literature more than one underlying factor has been detected, the structure of the instrument is recognized as essentially one-dimensional, which is why this is the recommended mode of use [57]. The analyses revealed good internal consistency (Cronbach’s *alpha* = 88.5).

#### Social support

To estimate the size of the social support network, the question “Approximately, how many close friends or relatives do you have? (people with whom you get along and can talk to about everything that is happening)” was used. This item is part of the MOS social support survey [58], also widely used in Spanish and in the Chilean context (e.g., [59]).

### Data Analysis Strategy

First, the factorial structure of the instruments was tested, applying confirmatory factor analysis (CFA) using JASP software version 0.11.1 (extraction methods used: maximum likelihood estimation, VARIMAX rotation), and the following indices were used for goodness of fit: *χ*^*2*^*/df*, RMSEA, SRMR, CFI, and TLI.

Second, along with previous analyses, the Harman test was applied to estimate possible common-method variance effects for all items in the study [60]. Third, once all the above were performed, the bivariate Pearson correlation was used to test the relationship between variables. Finally, to contrast the moderation hypothesis, Hayes’s macro [61] was applied using IBM SPSS 24 software.

## Results

In order to test the factor structure of the instruments, two CFAs was performed: the first analyzed the empirical structure of the data on the instruments of job insecurity along with the general health scale (N = 394; which corresponds to the participants who completed this scale, minus nine subjects self-categorized as “other occupational status”, i.e., the workers with full and part-time contracts and independent workers were considered in the analysis); the second model contains the financial threat scale along with the general mental health scale for the total sample. In both models the two latent variables freely covariate. As seen in Table 1, the goodness of fit indices used (*χ*^*2*^*/df*, SRMR, CFI, TLI) show a general good fit for the structure of the instruments, with the exception of RMSEA, which is acceptable. In the case of *χ2 / df* ratio, the result is very sensitive to large sample sizes, as is the case of the present study. However, according to the combination of adjustment index criteria established by [62], it can be argued that there is an adequate adjustment of the factorial structure of the instruments after ruling out one of the six items on the financial threat scale (How much do you think about your financial situation?), and two items on the general mental health scale (Have you felt that you are playing a useful role in life? and, Have you felt able to make decisions?), whose modification indexes indicated that its removal would allow a better adjustment of the data to the theoretical structure, which could be due to some level of redundancy in item content. The elimination of these items complies with the recommendation of not exceeding the twenty percent of them [63].

**Table 1.** Results of the CFA for the instruments.

| Model | $\chi^2$ | Df | $\chi^2/df$ ratio | SRMR | RMSEA | CFI | TLI |
| --- | --- | --- | --- | --- | --- | --- | --- |
| Work Insecurity (4 items) –<br>General Mental Health (12 items) | 442.290 | 103 | 4.300 | .068 | .092 | .868 | .847 |
| Work Insecurity (4 items) – General<br>Mental Health (10 items; eliminating 3<br>and 4) | 261.378 | 76 | 3.439 | .055 | .079 | .918 | .902 |
| Financial Threat (6 items) – General<br>Mental Health (12 items) | 724.734 | 134 | 5.408 | .069 | .099 | .864 | .844 |
| Financial Threat (5 items, eliminating 5)<br>– General Mental Health (10 items;<br>eliminating 3 and 4) | 301.762 | 89 | 3.391 | .053 | .073 | .939 | .929 |

Note: $N = 591$ ; SRMR = Standardized Root Mean Residual; RMSEA = Root Mean Square Error of Approximation; CFI = Comparative Fit Index; TLI = Tucker-Lewis Index.

To control the effect of common method variance (CMV), strategies were taken *a priori* and *post hoc*. Regarding the *a priori* strategy, it should be borne in mind that the scale formats for the different variables analyzed were different, which prevents the effects derived from CMV [64]. Regarding the *post hoc* strategy, we carried out the Harman single factor test [65]. This is the most widely used method to examine the pernicious presence of CMV [60, 66]. The results showed that this first single factor explains 24.43% of the variance, considerably less than the 50% from common variance; thus, CMV does not seem to affect correlations among the studied variables. Furthermore, Table 2 shows the descriptive statistics of the variables included in this study, as well as the respective Cronbach’s *Alpha* and McDonald’s *Omega*, which were reasonable (Table 2).

**Table 2.** Cronbach’s Alpha, McDonald’s Omega, Means, standards deviation and correlations.

| | $\alpha$ | $\omega$ | Mean | SD | Work Insecurity | Financial Threat | Social Support Network | General Mental Health |
| --- | --- | --- | --- | --- | --- | --- | --- | --- |
| Work Insecurity | .886 | .892 | 3.04 | 1.48 | 1 | .576** | -.074 | -.199** |
| Financial Threat | .911 | .914 | 3.04 | 1,10 |  | 1 | -.111** | -.343** |
| Social Support Network | --- | --- | 8.14 | 8,25 |  |  | 1 | .198** |
| General Mental Health | .875 | .877 | 2.93 | .55 |  |  |  | 1 |

Note: \* $p < .05$ \*\* $p < .01$ .

In order to test H1 and H2 bivariate correlations were run. As Table 2 shows, all of the study variable were significantly correlated, and particularly mental health was negatively related to both perceived work insecurity (*r* = -2.14, *p* < 0.05) and financial threat (*r* = -.335, *p* < 0.05), so H1 and H2 were accepted.

In order to test H3, a moderation analysis of social support network in the relationship between dependents and independents variables was carried out, controlling the effect of the variables sex, age and educational level (Table 3). From here, all the analyzes that account for direct effects and moderations use the scores of the independent variables transformed into z-scores. Results showed that the overall model significantly explained general mental health (*F*(6, 387) = 8.56, *p* < .001, *R*^2^ = .12). As shown in Table 3, both work insecurity and social support network predicted general mental health, and the interaction (moderation) between them contributed significantly to this explanation (*F*(1, 387) = 10.40, *p* = .001, *R*^2^ = .024), supporting H3.

**Table 3.** Moderation model of social support on work insecurity and general mental health.

| Variables | Coefficient | t | p | LI | LS |
| --- | --- | --- | --- | --- | --- |
| Constant | 2.439 | 12.459 | .000 | 2.054 | 2.824 |
| SSN | .114 | 4.392 | .000 | .063 | .165 |
| WI | -.082 | -3.379 | .001 | -.130 | -.034 |
| SSNxWI | .100 | 3.225 | .001 | .039 | .162 |
| SX | .096 | 1.771 | .077 | -.011 | .203 |
| AG | .004 | 1.466 | .144 | -.001 | .009 |
| EL | .060 | 1.696 | .091 | -.010 | .130 |

Note: 95% confidence level. SSN = Social Support Network, WI = Work Insecurity, SX = Sex, AG = Age, EL = Educational Level.

Figure 1 shows the effect of work insecurity over general mental health, in presence of social support network. It is possible to see that when social support network is high, the effect of work insecurity on general mental health is not significant.

**Figure 1.**
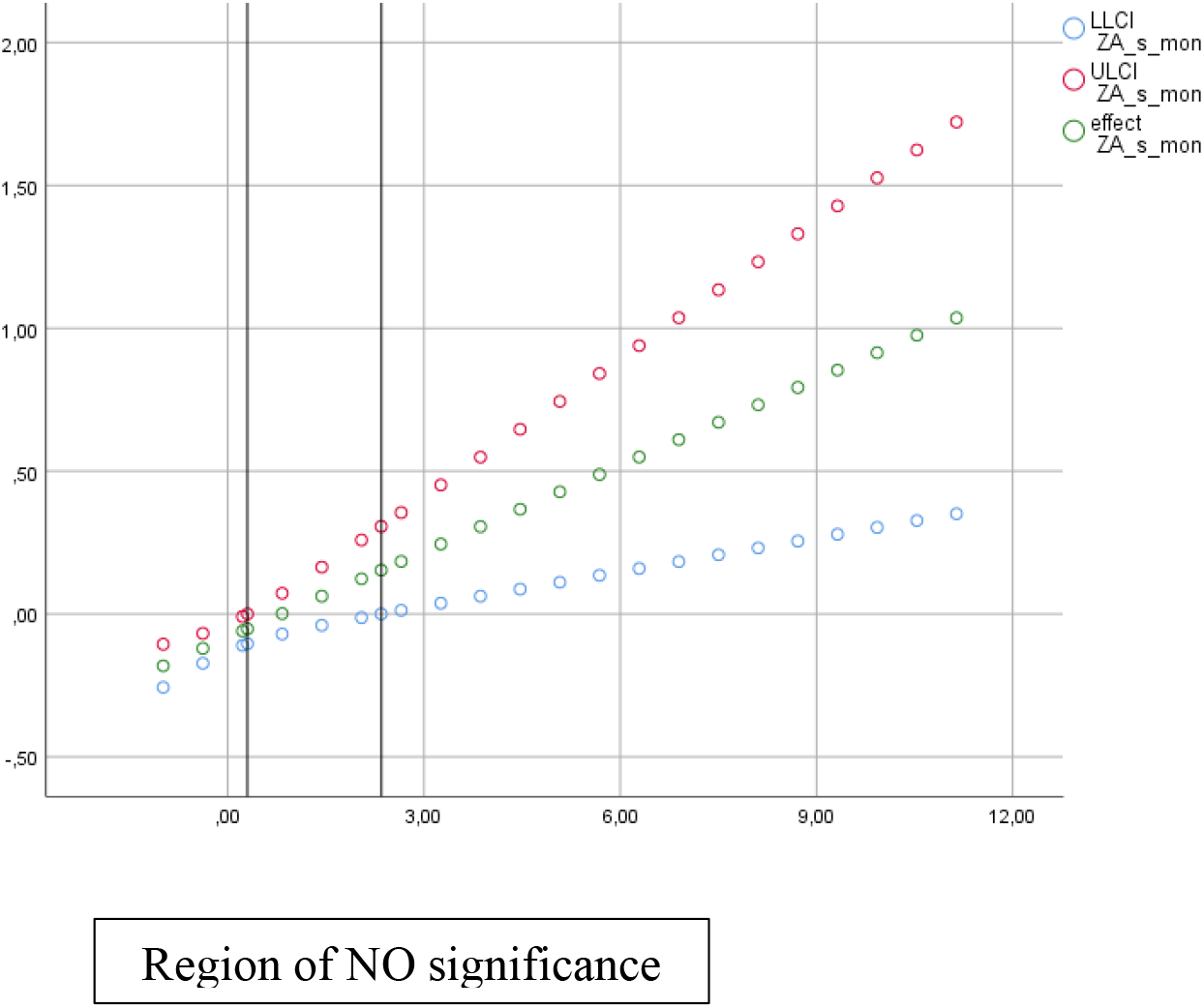
Conditional effect of work insecurity on perceived general mental health.

Next, we tested the moderation of social support network in the relationship between financial threat as independent variable, and general mental health as dependent variable, controlling the effect of the variables sex, age, and educational level. Results showed that the overall model significantly explained general mental health (*F*(6, 583) = 24.22, *p* < .001, *R*^2^ = .20). As shown in Table 3, both financial threat and social support predicted general mental health and, supporting H3, the interaction (moderation) between them contributed significantly to this explanation (*F*(1, 583) = 5.55, *p* = .018, *R*^2^ = .008).

**Table 4.** Moderation model of social support on financial threat and general mental health.

| Variables | Coefficient | T | P | LI | LS |
| --- | --- | --- | --- | --- | --- |
| Constant | 2.354 | 20.453 | .000 | 2.128 | 2.580 |
| SSN | .096 | 4.217 | .000 | .052 | .141 |
| FT | -.161 | -7.726 | .000 | -.202 | -.120 |
| SSNxFT | .042 | 2.357 | .018 | .007 | .078 |
| SX | .100 | 2.132 | .033 | .008 | .192 |
| AG | .008 | 4.629 | .000 | .004 | .011 |
| EL | .040 | 1.764 | .078 | -.004 | .084 |

Note: 95% confidence level. SSN = Social Support Network, FT = Financial Threat, SX = Sex, AG = Age, EL = Educational Level.

As Figure 2 shows, the effect of financial threat on perceived general health is buffered by social support network, that is, when social support network is high, the effect of financial threat on perceived general mental health is not significant.

**Figure 2.**
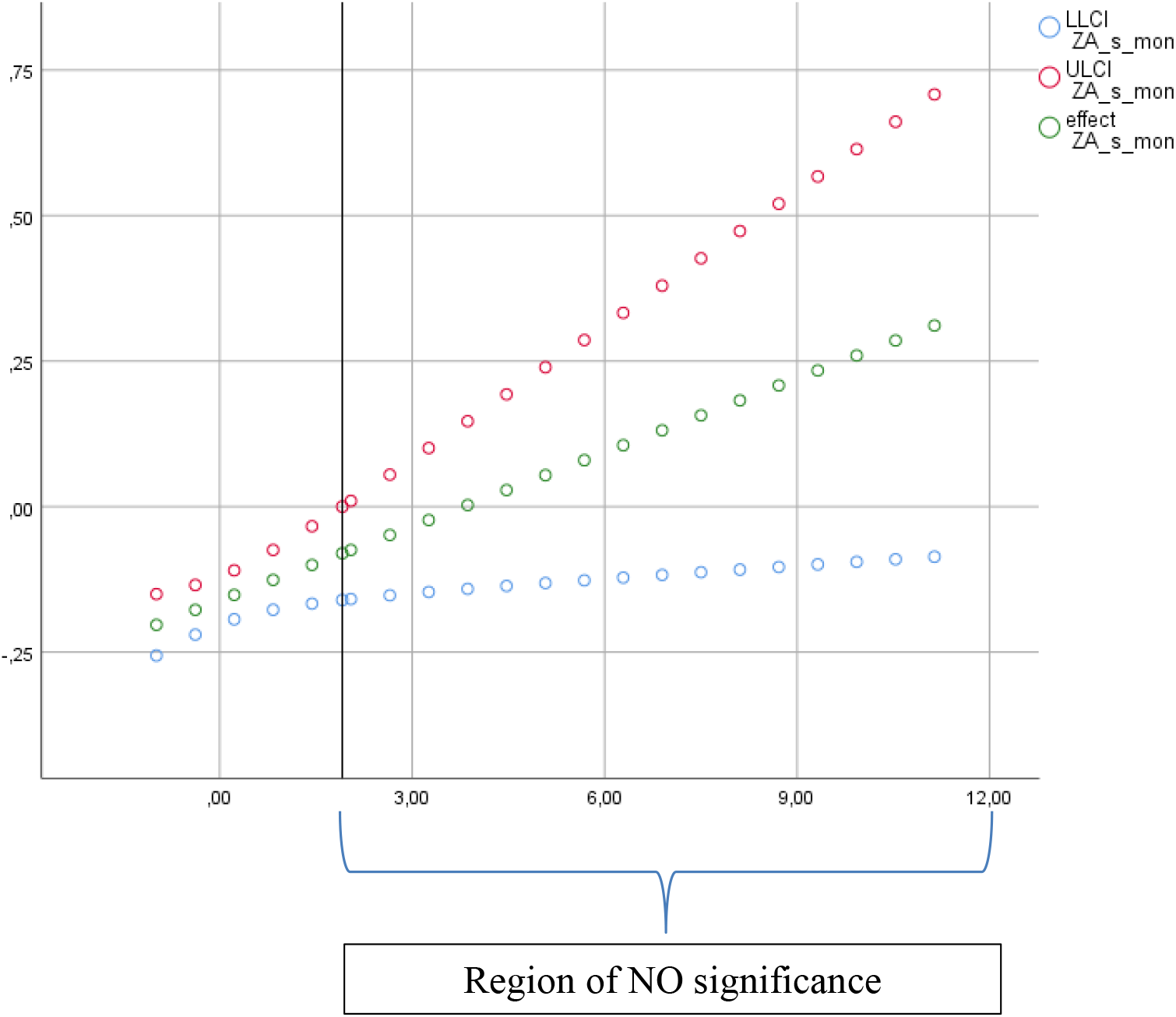
Conditional effect of financial threat on general mental health.

## Discussion

As the months pass, the effects of ‘Coronanomics’ [1] are amplifying the devastating effects on health that COVID-19 is causing. By the mid-July, Chile was the eighth country of the world in number of deaths per number of inhabitants with a rate of 45.40 deaths per 100,000 inhabitants [67]. In April, the World Bank estimated that Chile’s economic activity could shrink up to 3.3% in 2020, accompanied by a significant reduction in trade flows, consumption and the price of raw materials, especially copper, which will result in increased unemployment and poverty rates [68]. The data from our study indicate that people already perceived this impact during first stage of the pandemic through job insecurity and financial threat, and that these perceptions related negatively to their overall mental health, as is discussed next.

First, the measurement of global job insecurity [17, 20] indicates that this issue is perceived as a threat in Chile in the context of the COVID-19 pandemic, although the overall mean value is not high. It is possible that the education level of the sample, with a predominance of university graduates, is related to a perception of greater employability, which helps to relativize the perception of job insecurity and its negative psychological consequences [69, 70]. However, there are studies that have not identified a buffer effect of employability on the positive relation between job insecurity and the deterioration of psychological well-being [71].

The results show a statistically significant relation between job insecurity and mental health, which agrees with the evidence obtained in other socioeconomic contexts [19, 28-29] and makes it possible to verify that this relation is consistent across countries and cultures. The current situation brought about by COVID-19 fulfills two of the conditions that characterize job insecurity according to [23] –lower control and lower volition– since the impact generated by the pandemic has been sudden and uncontrollable for governments and for organizations and workers alike. Although the potential effect of the third of these – longer duration [23] –will possibly be proven later on, it must be remembered that Chile had undergone the effects of the “social uprising” in October and November 2019 and its consequences for economic and social activities, which could also contribute to increase the perception of job insecurity as the two effects are now combined. Accordingly, it is important to warn of the potential development of post-traumatic stress and the declining on mental health in the medium and long term [72, 73] if the adverse social and employment conditions persist or even become more acute, especially in the most vulnerable groups and sectors with greater psychosocial and material needs.

Second, in the same direction, the outcomes show a perception of financial threat at values slightly above the midpoint of the scale. The relatively rapid measure approved by the Chilean Senate and Congress to make unemployment insurance more flexible due to the Covid-19 emergency (March 31), which extended coverage for the most vulnerable sectors, could contribute to control the perception of immediate financial risk, even if it is on a relatively small scale. However, the results show a statistically significant relation with mental health, which agrees with previous evidence after the effects of large economic recessions [7, 35, 37, 41]. This conclusion is relevant because it is consistent with the increasing body of evidence (e.g. [40]) that economic insecurity is experienced the same regardless of income distribution, with identical effect of psychological distress.

In previous crisis brought about by epidemics and pandemics, the effects of financial threat are detected both during the quarantine period and, especially, in the medium and long term [74, 75]. As a result, if the outcomes of our study show that the mean levels of financial threat are already associated with declining mental health in the first stage of the COVID-19 pandemic, it is foreseeable that when the current situation is prolonged, the economic stressors will increase, with the potential consequent deterioration in well-being and psychological health. Prior research is consistent in showing that the negative and adverse effects of repeated financial threat [76], associated with both job insecurity and unemployment, are multiple and severe [77] for people and their personal and social environments – affecting in particular social relations and family functioning [78]; therefore, the outcomes obtained in this study can serve to warn of the foreseeable consequences for mental health in an employment and economic situation that may continue over time.

The results show that the perceptions of job insecurity and financial threat are significantly related to each other, which agrees with what has been found in previous studies [8, 44] in situations of recent large-scale economic crises. In addition, both are associated with a decline in perceived mental health, which is also consistent with prior results (e.g. [8, 40, 77]). However, our data indicate that the financial threat is slightly more negatively related to mental health (*r* = -.335) than to job insecurity (*r* = -.214). These results agree in part with the prior evidence [30], but not with the results obtained by [8] in the context of the Great Recession of 2008 in European countries, where they verified that job insecurity and financial worries affected mental health the same. Nor are they in line with those obtained in Sweden by [79], who confirmed job dependency due to economic issues did not mitigate the relation between job insecurity and psychological well-being. These discrepancies with some prior results may be because in Chile social protection systems are much less robust than those generally available in European Union countries with welfare states [8], and in particular in Sweden [79], and they may be close to those that predominate in the US [30]. Thus, in Chile, where the population generally carries high debt levels and a fragile unemployment insurance system, people who view their financial situation with concern are more job-dependent and react more negatively to the threat, which is reflected in their mental health. In this sense, we argue that the moderating effect of the social support network will be less if the person who works is responsible for a larger number of people. In order to analyze it, we performed a hierarchical regression, segmenting the file into two groups (1 or fewer dependents = group 1; two or more dependents = group 2, *ns* = 261 and 131 respectively) to verify the direct effects of financial threat, social support network and their interaction on overall mental health. In line with the previous reasoning, the results showed that in both groups direct significant regression weights of financial threat and social support network appear on overall mental health (*β*s > .108, *p*s < .005), but that the interaction is only related to a lower level of damage to mental health in the group of up to one dependent (*β*s = .087 and .000, *p*s = .002 and .996, for groups 1 and 2, respectively). Additionally, the percentage of variance explained by these models is greater in group 2 than in group 1 (*R*^2^ = .166 and .123, respectively), which is why these variables to a large extent explain the worsening of overall mental health in the group with more dependents. Prior results [40] also show that the negative effects of economic insecurity are greater in men, who for socioeconomic and cultural reasons continue to be the main providers in families, as can also occur in Chile.

Finally, the study results indicate a moderate buffer effect of perceived social support relative to the size of the social network. Thus, in relation to job insecurity and financial threat, the network size mitigates the association of both with the decline in perceived mental health. These results are similar to those obtained in previous studies in relation to both constructs [12-16, 49], and reinforce the value of social support as a resource to cope with stressful experiences beyond specific situations and contexts. Although the network size of the participants is not high (mean 8.13 average), what seems important is the perceived effectiveness of this network to manage job insecurity and financial threat. When asking for the number of people who form their support network, we were investigating not only the size of the social network, but also the cohesion of the network and the type of close relations (i.e., primary and strong bonds) that people establish in them. Both factors directly affect the reception of various types of social support [80], so that the participants seem to perceive that the social network is a powerful resource to cope with financial stress and job insecurity. In addition to this direct influence, the perception of the support network may also expand through the experience of trust, where its members increase their social capital through their own contacts.

### Practical implications

The outcomes obtained have direct practical implications related to the need to implement psychological support measures for workers and the unemployed in order to equip them with coping strategies to manage uncertainty and stress. These psychosocial supports should be available in organizations and primary care centers so they can be accessible and have a restraining effect on the most serious mental health consequences in the medium and long term [72].

Second, the conclusions drawn here should serve as an alert to the risk that the double perception of job insecurity and financial threat can generate by increasing the psychological pressures of productivity, one of the first consequences that has already been detected since the pandemic began [81]. The demands, explicit or implicit, that people respond to the challenges caused by the COVID-19 crisis with work overload, increasing their working days (in person or remotely), reducing or giving up their days off or vacation days, and accepting abusive working conditions, can also increase the perception of qualitative job insecurity [21], lower perceived control and the resulting negative effects on mental and physical health [82]. Thus, it seems urgent to regulate and monitor compliance with working conditions during the COVID-19 crisis and the call for “the new normal”, as well as to guarantee the rights of workers and the unemployed, to maintain adequate levels of well-being and physical and mental health, and to reduce other negative psychosocial consequences such as the work-family conflict.

Third, the results also have important implications for the design of strategies and psychosocial interventions that can strengthen social networks and the potential social support they involve. Given the strong bond between social relations, support and mental health [46-47], actions aimed at reinforcing interpersonal relations and social networks through, for example, support groups, community initiatives or networks of people who share similar characteristics (studies, profession, etc.) [83] can be highly effective at equipping those who experience financial threat and job insecurity with social coping and resilience resources that increase personal resources. Despite being subjective perceptions, financial and job insecurity have objective components and, consequently, are not an *individual* but a collective problem; therefore, the possible tools to confront them will also need to be collective and social.

### Limitations and Future Research

Although the result of Harman’s test indicates the absence of common-method variance effects, the cross-sectional nature of the design only allows us to identify relations between the variables, although these relations were statistically significant. However, despite this limitation, the importance of having preliminary data on the disruptive impact caused by the scale and speed of the global pandemic in Chile justifies the urgency of obtaining an initial approach of the consequences for mental health of work-related variables. Indeed occupational health, job insecurity and job precariousness are three of the ten key areas for research and practice in WOP due to the impact on them by COVID-19 as identified by experts (Rudolph et al., 2020); thus, this study makes an important contribution to knowledge in this context, but it must be broadened or complemented.

In this regard, our aim is to continue collecting data on these variables so that longitudinal studies can establish causal relations among them, and to learn with greater precision the long-term effects and perceptions of financial threat and psychological insecurity on mental health.

Third, the participants in our study did not form part of a probabilistic sample, so that the outcomes cannot be generalized to the Chilean population. Also in this case, future studies will have to try to access representative samples, or concentrate groups of workers, specific people or contexts (concrete professions, young or older workers or vulnerable groups, etc.) in order to most accurately identify the differential effects of ‘Coronanomics’ [1] in each setting.

Finally, this study has only considered the variables mentioned as moderating antecedents and consequents, but we are aware that in the experiences and the outcomes that people are living during this economic and health crisis other personal, family, organizational and social variables are involved that interact in a complex way, which is reflected in the percentage of variance explained by the contrasted models not exceeding 20%. Their inclusion in future studies will be needed to understand the real impacts and to design strategies and interventions that can prevent, reduce or alleviate the effects for the well-being, health and safety of people and their families as well as for the functioning and effectiveness of organizations.

## Data Availability

The data corresponding to this study is available on Open Science Framework (OSF) website

https://osf.io/p7xdf/

## Data availability

The data corresponding to this study is available on Open Science Framework (OSF) website (https://osf.io/p7xdf/) [84].

## Acknowledgments

This research has been supported by project CONICYT MEC CON80190085 (Chile), and by Research Directorate (DIUFRO) of the Universidad de La Frontera (Chile).

## Notes

### Competing Interest Statement

The authors have declared no competing interest.

### Author Declarations

The project underwent assessment by the Ethics and Bioethics Committee of the Universidad de Concepcion (CEBB 650-2020, March 2020), and all the participants were asked to sign an informed consent.

## References

1. Eichengreen B. Coronanomics 101: which policy tools will contain the economic threat of COVID-19? World Economic Forum, 2020 Mar 12. Available from: https://www.weforum.org/agenda/2020/03/coronavirus-economics/

2. Barua S. Understanding Coronanomics: The economic implications of the coronavirus (COVID-19) pandemic. MPRA Paper No. 99693, posted 2020 Apr 20. Available from: https://papers.ssrn.com/sol3/papers.cfm?abstract_id=3566477

3. Burgard SA, Brand JE, House JS. Perceived job insecurity and worker health in the United States. Soc Sci Med. 2009; 69: 777–785.

4. De Witte H. On the scarring effects of job insecurity (and how they can be explained). Scan J Work Environ Health. 2016; 42: 99–102.

5. Lübke C. How self-perceived job insecurity affects health: Evidence from an age-differentiated mediation analysis. Econ. Ind. Dem. 2019 Forthcoming.

6. Kiely KM, Leach LS, Olesen SC, Butterworth P. How financial hardship is associated with the onset of mental health problems over time. Soc Psych & Psych Epidem: Int J Res Soc Gen Epidem Ment Health Serv. 2015; 50: 909–918.

7. Marjanovic Z, Greenglass ER, Fiksenbaum L, Bell CM. Psychometric evaluation of the Financial Threat Scale (FTS) in the context of the great recession. J Econ Psych. 2013; 36: 1–10.

8. Rajani NB, Giannakopoulos G, Filippidis FT. Job insecurity, financial difficulties and mental health in Europe. Occup Med. 2016; 66: 681–683.

9. László KD, Pikhart H, Kopp MS, Bobak M, Pajak A, Malyutina S, Salavecz G, Marmot M. Job insecurity and health: A study of 16 European countries. Soc Sci Med. 2010; 70: 867–874.

10. Yip PSF, Yang KCT, Ip BYT, Law YW, Watson R. Financial debt and suicide in Hong Kong SAR. J Appl Soc Psychol. 2007; 37: 2788–2799.

11. Mauno S, Cheng T, Lim V. The far-reaching consequences of job insecurity: A review on family-related outcomes. Marr Family Rev. 2017; 53: 717–743.

12. Näswall K, Sverke M, Hellgren J. The moderating effects of work-based and non-work based support on the relation between job insecurity and subsequent strain. SA J Indus Psychol. 2005; 31: 57–64.

13. Snow DL, Swan SC, Raghavan C, Connell CM, Klein I. The relationship of work stressors, coping and social support to psychological symptoms among female secretarial employees. Work Stress. 2003; 17: 241–263.

14. Lim VKG. Job insecurity and its outcomes: Moderating effects of work-based and nonwork-based social support. Hum Relat. 1996; 49: 171–194.

15. Åslund C, Larm P, Starrin B, Nilsson KW. The buffering effect of tangible social support on financial stress: Influence on psychological well-being and psycho-somatic symptoms in a large sample of the adult general population. Int J Equity Health. 2014; 13: 85–93.

16. Whelan CT. The role of social support in mediating the psychological consequences of economic stress. Soc Health Illness, 1993; 15: 86-101.

17. De Witte H. Job insecurity: Review of the international literature on definitions, prevalence, antecedents and consequences. SA J Ind Psychol. 2005; 31: 1–6.

18. Sverke M., Hellgren J. The nature of job insecurity: Understanding employment uncertainty on the brink of a new millennium. Appl Psychol: An Int Rev. 2002; 51: 23–42.

19. Sverke M, Hellgren J, Näswall K. No security: A meta-analysis and review of job insecurity and its consequences. J Occup Health Psychol. 2002; 7: 242–264.

20. Vander Elst T, De Witte H, De Cuyper N. The Job Insecurity Scale: A psychometric evaluation across five European countries. Eur J Work Org Psychol. 2014; 23: 364–380.

21. Hellgren J, Sverke M, Isaksson K. A two-dimensional approach to job insecurity: Consequences for employee attitudes and well-being. Eur J Work Org Psychol. 1999; 8: 179–195.

22. Gallie D, Felstead A, Green F, Inanc H. The hidden face of job insecurity. Work Employ Soc. 2017; 31: 36–53.

23. Shoss MK. Job insecurity: An integrative review and agenda for future research. J Manage. 2019; 43:1911-1939.

24. De Cuyper N, Piccoli B, Fontinha R, De Witte H. Job insecurity, employability and satisfaction among temporary and permanent employees in post-crisis Europe. Econ Ind Dem. 2019; 40: 173–192.

25. De Witte H, Vander Elst T, De Cuyper N. Job Insecurity, Health and Well-Being. In Vuori J, Blonk R, Price RH, editors. Sustainable Working Lives, Aligning Perspectives on Health, Safety and Well-Being. Cham: Springer; 2015. pp. 109–128

26. Jiang L, Lavaysse LM. Cognitive and affective job insecurity: A meta-analysis and a primary study. J Manage. 2018; 44: 2307–2342.

27. De Witte H. Arbeidsethos en jobonzekerheid: Meting en gevolgen voor welzijn, tevredenheid en inzet op het werk [Work ethic and job insecurity: Assessment and consequences for wellbeing, satisfaction and performance at work]. In Bouwen R, De Witte k, De Witte h, Taillieu T, editors. Van groep naar gemeenschap [From group to community]. Liber Amicorum Prof. Dr. Leo Lagrou. Leuven, Belgium: Garant; 2000. pp. 325–350.

28. Cheng G. Chan D. Who Suffers More from Job Insecurity? A Meta-Analytic Review. Appl Psychol: An Int Rev. 2008; 57: 272–303.

29. Llosa JA, Menéndez-Espina S, Agulló-Tomás E, Rodríguez-Suárez J. Job insecurity and mental health: A meta-analytical review of the consequences of precarious work in clinical disorders. Anales Psicol 2018; 34: 211-223.

30. Greenhalgh L, Rosenblatt Z. Job insecurity: Toward conceptual clarity. The Acad Manage Rev. 1984; 9: 438–448.

31. Caroli E, Godard M. Does job insecurity deteriorate health? Health Econo. 2016; 25: 131–147.

32. Virtanen M, et al. Perceived job insecurity as a risk factor for incident coronary heart disease: systematic review and meta-analysis. BMJ, 2013; 347:f4746.

33. Otterbach S, Sousa-Poza A. Job Insecurity, Employability, and Health: An Analysis for Germany across Generations. Applied Econo. 2016; 48: 1303–1316.

34. Lozza E, Castiglioni C, Bonanomi A. The effects of changes in job insecurity on daily consumption and major life decisions. Econo Ind Demo. 2017. Forthcoming.

35. Fiksenbaum L, Marjanovic Z, Greenglass E. Financial threat and individuals’ willingness to change financial behavior. R Behav Finan. 2017; 9: 128–147.

36. Giannetti C, Madia M, Moretti L. Job insecurity and financial distress. Applied Finan Econo. 2014; 24: 219–233.

37. Marjanovic Z, Greenglass ER, Fiksenbaum L, De Witte H, Garcia-Santos F, Buchwald P, Peiró JM, Mañas MA. Evaluation of the financial threat scale (FTS) in four European, non-student samples. J Behav Exp Econo. 2015; 55: 72–80.

38. Blázquez M, Budría S, Moro-Egido AI. Job Insecurity, Debt Burdens and Individual Health. IZA Discussion Papers, No. 12663. Bonn: Institute of Labor Economics (IZA); 2019.

39. Drentea P. Age, debt and anxiety. J Health Soc Behav. 2000; 41: 437–450.

40. Kopasker D, Montagna C, Bender K. Economic Insecurity as a Socioeconomic Determinant of Mental Health. SSM - Popul Health. 2018; 6: 184–194.

41. Netemeyer RG, Warmath D, Fernandes D, Lynch JG. How am I doing? Perceived financial well-being, its potential antecedents, and its relation to overall well-being. J Cons Res. 2018; 45: 68–89.

42. de Bruijn EJ, Antonides G. Determinants of financial worry and rumination. J. Econ Psychol. 2020; 76: [102233].

43. Fiksenbaum L, Marjanovic Z, Greenglass E, Garcia-Santos F. Impact of Economic Hardship and Financial Threat on Suicide Ideation and Confusion. J. Psychol. 2017; 151: 477–495.

44. Lam J, Wen F, Phyllis M. Is Insecurity Worse for Well-being in Turbulent Times? Mental Health in Context. Soc Men Health. 2014; 4: 55–73.

45. Glavin P. The impact of job insecurity and job degradation on the sense of personal control. Work Occup. 2013; 40: 115–142.

46. House JS, Landis KR, Umberson D. Social relationships and health. Science. 1988; 241: 540–545.

47. Uchino BN. Social support and health: A review of physiological processes potentially underlying links to disease outcomes. J Behav Med. 2006; 29: 377–387.

48. Cohen S, Wills TA. Stress, social support, and the buffering hypothesis. Psychol Bull. 1985; 98: 310–357.

49. Cheng T, Mauno S, Lee C. Do job control, support, and optimism help job insecure employees? A three-wave study of buffering effects on job satisfaction, vigor and work-family enrichment. Soc Indic Res. 2014; 118:1269-1291.

50. Matud MP. Gender differences in stress and coping styles. Pers Indiv Differ. 2004; 37: 1401–1415.

51. Menéndez-Espina S, Llosa JA, Agulló-Tomás E, Rodríguez-Suárez J, Sáiz-Villar R, Lahseras-Díez HF. Job insecurity and mental health: The moderating role of coping strategies from a gender perspective. Front Psychol. 2019; 10:286.

52. Giunchi M, Vonthron A-M, Ghislieri C. Perceived job insecurity and sustainable wellbeing: do coping strategies help? Sustainability 2019; 11: 784.

53. Peirce RS, Frone MR, Russell M, Cooper ML. Financial stress, social support, and alcohol involvement: A longitudinal test of the buffering hypothesis in a general population survey. Health Psychol. 1996; 15: 38–47.

54. Goldberg DP, Williams P. A user’s guide to the General Health Questionnaire. Windsor: NFERNelson; 1988.

55. Garmendia ML. Análisis factorial: una aplicación en el cuestionario de salud general de Goldberg, versión de 12 preguntas [Factorial analysis: an application in Goldberg’s general health questionnaire, 12-item versión]. Rev Chil Salud Pub. 2007; 11: 57–65.

56. Rivas-Diez R, del Pilar Sanchez-Lopez M. Psychometric properties of the general health questionnaire (GHQ-12) in chilean female population/Propiedades psicométricas del cuestionario de salud general (GHQ-12) en población femenina chilena. Rev Argen Clin. Psicol. 2014; 23: 251–260.

57. Gnambs T, Staufenbiel T. 2018. The structure of the General Health Questionnaire (GHQ-12): two meta-analytic factor analyses. Health Psychol Rev. 2018; 12:179-194.

58. Sherbourne C, Stewart A. The MOS Social Support Survey. Soc Sci Med. 1991; 32: 705–714.

59. Poblete F, Glasinovic A, Sapag J, Barticevic N, Arenas A, Padilla O. Apoyo social y salud cardiovascular: adaptación de una escala de apoyo social en pacientes hipertensos y diabéticos en la atención primaria chilena [Social support and cardiovascular health: Adaptation of a scale of social support in hypertensive and diabetic patients in Chilean primary care]. Aten Prim. 2015; 47: 523–531.

60. Podsakoff PM, MacKenzie SB, Lee J, Podsakoff N. Common method biases in behavioral research: A critical review of the literature and recommended remedies. J App Psychol. 2003; 88: 879–903.

61. Hayes A. Methodology in the social sciences. Introduction to mediation, moderation, and conditional process analysis: A regression-based approach. New York: Guilford Press; 2013.

62. Hu L, Bentler P. Cutoff criteria for fit indexes in covariance structure analysis: Conventional criteria versus new alternatives. Struct Equat Model: A Multidisc J 1999; 6: 1-55.

63. Kline RB. Principles and practice of structural equation modeling; 2nd edition. New York: Guilford Press; 2005.

64. Podsakoff PM, MacKenzie SB, Podsakoff NP. Sources of method bias in social science research and recommendations on how to control it. Annual Rev. Psychol. 2012; 63: 539–569.

65. Chang SJ, van Witteloostuijn A, Eden L. From the editors: Common method variance in international business research. J Inter Bus Stud. 2010; 41: 178–184.

66. Tehseen S, Ramayah T, Sajilan S. Testing and controlling for common method variance: A review of available methods. J Manage Sci. 2017; 4: 142–168.

67. Johns Hopkins Coronavirus Resource Center (2020). Mortality Analysis. 2020 July 17. Available from: https://coronavirus.jhu.edu/data/mortality

68. Menz E. Impacto del COVID-19 en la economía de América Latina y Chile. FLACSO-Chile; 2020 May 28. Available from: http://www.flacsochile.org/slider/articulo-impacto-del-covid-19-en-la-economia-de-america-latina-y-chile/

69. De Cuyper N, Mäkikangas A, Kinnunen U, Mauno S, De Witte H. Cross-lagged associations between perceived external employability, job insecurity, and exhaustion: Testing gain and loss spirals according to the Conservation of Resources Theory. J Org Behav. 2012; 36: 770–788.

70. Green F. Unpacking the misery multiplier: How employability modifies the impacts of unemployment and job insecurity on life satisfaction and mental health. J Health Econ. 2011; 30: 265–276.

71. Kirves K, De Cuyper N, Kinnunen U, Nätti J. Perceived job insecurity and perceived employability in relation to temporary and permanent workers’ psychological symptoms: A two samples study. Intern Arch Occup Environ Health. 2011; 84: 899–909.

72. Pfefferbaum B, North, CS. Mental Health and the COVID-19 Pandemic. The New Engl J Med; April 13, 2020. Available from: https://www.nejm.org/doi/full/10.1056/NEJMp2008017

73. Torales J, O’Higgins M, Castaldelli-Maia JM, & Ventriglio A. The outbreak of COVID-19 coronavirus and its impact on global mental health. Intern J Soc Psych. 2020; 66: 317–320.

74. Brooks SK, Webster RK, Smith LE, Woodland L, Wessely S, Greenberg N, Rubin, GJ. The psychological impact of quarantine and how to reduce it: Rapid review of the evidence. Lancet. 2020; 395: 912–20.

75. Jeong H, Yim HW, Song Y-J, Ki M, Min J-A, Cho J, Chae J-H. Mental health status of people isolated due to Middle East respiratory syndrome. Epidem Health 2016; 38: e2016048.

76. Watson B, Osberg L. Healing and/or breaking? The mental health implications of repeated economic insecurity. Soc. Sci Med. 2017; 188: 119–127.

77. Klehe U-C, Van Vianen Aem, Zikic J. Coping with economic stress: Introduction to the special issue. J Org Behav. 2012; 33: 745–751.

78. Probst TM. Economic stressors. In Barling J, Kelloway EK, Frone, MRs; editors. Handbook of Work Stress. London: Sage; 2005. pp. 267–297.

79. Richter A, Näswall K, Bernhard-Oettel C, Sverke M. Job insecurity and well-being: The moderating role of job dependence. Euro J Work Org Psychol. 2014; 26: 816–829.

80. Thoits PA. Mechanisms Linking Social Ties and Support to Physical and Mental Health. J Health Soc Behav. 2011; 52: 145–161.

81. Mukhtar S. Psychological health during the coronavirus disease 2019 pandemic outbreak. Int J Soc Psych. 2020; Forthcoming.

82. Vander Elst T, Richter A, Sverke M, Näswall K, De Cuyper N, De Witte H. Threat of losing valued job features: The role of perceived control in mediating the effect of qualitative job insecurity on job strain and psychological withdrawal. Work Stress. 2014; 28: 143–164.

83. Cohen S, Gottlieb BH, Underwood LG. Social relationships and health. In Cohen S, Underwood LG, Gottlieb BH, editors. Social support measurement and intervention: A guide for health and social scientists. Oxford: Oxford University Press; 2000. pp. 3–25.

84. Salgado S. Job Insecurity, Financial Threat and Mental Health in the COVID-19 Context: The Buffer Role of Perceived Social Support [Internet]. OSF; 2020. Available from: osf.io/p7xdf

